# The COVID-19 Early Detection in Doctors and Healthcare Workers (CEDiD) Study: study protocol for a prospective observational trial

**DOI:** 10.1101/2020.08.11.20172502

**Authors:** Alexander Zargaran, Dina Radenkovic, Iakovos Theodoulou, Andrea Paraboschi, Chelsea Trengrove, Gary Colville, Gill Arbane, Kariem El-Boghdadly, Gaia Nebbia, Rocio Martinez-Nunez, Anne Greenough

## Abstract

**Background:** The global COVID-19 pandemic has caused worldwide disruption with its exponential spread mandating national and international lockdown measures. Hospital-associated transmission has been identified as a major factor in the perpetuation of COVID-19, with healthcare workers at high-risk of becoming infected with SARS-CoV-2 and representing important vectors for spread, but not routinely having their clinical observations monitored or being tested for SARS-CoV-2.

**Methods:** A single-center, prospective observational study of 60 healthcare workers will explore how many healthcare workers in high-risk areas develop COVID-19 infection over a thirty day period. High-risk areas are defined as COVID positive wards, the intensive care unit and the accident and emergency department. Healthcare workers (HCWs) will be recruited and have daily self-administered nasopharyngeal SARS-CoV-2 PCR tests. They will also be provided with a wearable medical device to measure their clinical observations during non-working hours, and be asked to complete a daily self-reported symptom questionnaire over the study period. Statistical analysis will assess the proportion of healthcare workers who develop COVID-19 infection as a primary objective, with secondary objectives exploring what symptoms are developed, time-to-event, and deviations in clinical observations.

**Discussion:** At present clinical observations, symptoms and SARS-Cov-2 PCR swabs are not routinely undertaken for healthcare workers. If the CEDiD (COVID-19 Early Detection in Doctors and Healthcare Workers) study is successful, it will provide useful information for workforce decisions in reducing hospital-associated transmission of COVID-19. The data will help in determining whether there are early warning signs for development of COVID-19 infections amongst healthcare workers and may contribute to evidence based advocating for more regular testing of healthcare workers’ observations, symptoms and COVID-19 status.

**Trial registration:** ClinicalTrials.gov, NCT04363489. Registered on 27^th^ July 2020

## Background

The global COVID-19 pandemic has caused exceptional pressures on health services worldwide, which in many cases have been overwhelmed by the number and severity of cases. Challenges spanning recognition, management and containment strategies being amongst the most important considerations for service providers. Whilst existing research predominantly focuses on recognition and management of COVID-19 in patients, reducing transmission of SARS-CoV-2 within a healthcare setting is vital. In the SARS 2002-03 outbreak in Toronto, hospital transmission was cited as a major factor perpetuating the spread of the disease (1). The estimated human-to-human hospital-associated transmission of COVID-19 in Wuhan was reported as 41% (2). Furthermore, public anxiety surrounding hospitals and the likelihood of contracting COVID-19 has unfortunately resulted in delayed presentations to hospital, with many treatable diseases progressing without timely medical interventions (3). The reopening of services in the aftermath of the two peaks of cases in the United Kingdom (UK) means that hospital-associated transmission must be mitigated in order to prevent the spread of COVID-19 and in anticipation of likely future waves.

International estimates of COVID-19 infection amongst HCWs range between 4-19% in Europe, the United States and China (4,5). HCWs are at high risk of both contracting and spreading COVID-19 given their frequent face-to-face interactions with patients who are COVID-19 positive. One study has reported that frontline HCWs have an adjusted-Hazard Ratio of a positive COVID-19 test of 11.6 (95% CI: 10.9 to 12.3) when compared to the general population, which decreases to 4.83 (95% CI: 3.99 to 5.85) when they have adequate PPE (6). A survey conducted by the Royal College of Physicians in March 2020 identified that 11% of doctors were taking time off work due being symptomatic with COVID-19 or having a symptomatic household member (7). In the context of the asymptomatic rate of infected individuals being approximately one in five (8), these numbers highlight that HCWs are at risk of both becoming infected with SARS-CoV-2 and spreading the infection. At present, aside from the newly introduced voluntary weekly staff testing, no concurrent strategies have been implemented to screen for COVID-19 in healthcare workers. With the variety of possible viral infections that are prevalent during the winter months, reliable strategies must be devised and implemented in order to identify COVID-19 infection in HCWs at the earliest possible stage to prevent the spread of the disease.

As a consequence, we have designed the “COVID-19 Early Diagnosis in Doctors and Healthcare Workers” (CEDID) Study, a cohort study of HCWs. The trial has been reviewed by The Health Research Authority and received permission to proceed with data collection. If the CEDiD Study is successful, it could pave the way for wider-scale testing and observation monitoring of HCWs, enable earlier detection of COVID-19, and help to prevent its spread within hospitals.

## Objectives

The primary objective of The CEDiD Study is to identify how many healthcare workers who work in high-risk COVID-19 areas develop the infection over the study period. Secondary objectives of the CEDiD Study are to explore the trend in clinical observations and daily self-reported symptoms of COVID-19, as well as to validate an algorithm for early detection of COVID-19 in healthcare workers.

## Methods

The CEDiD study (NCT04363489) is a prospective observational cohort study of 60 HCWs working in high-risk areas for COVID-19. This manuscript has been prepared in line with the Strengthening the Reporting of Observational Studies in Epidemiology (STROBE) statement (9). The STROBE Checklist (Additional file 1), and SPIRIT Checklist (Additional file 2 and Figure 1) for this study are attached.

**Figure 1:**
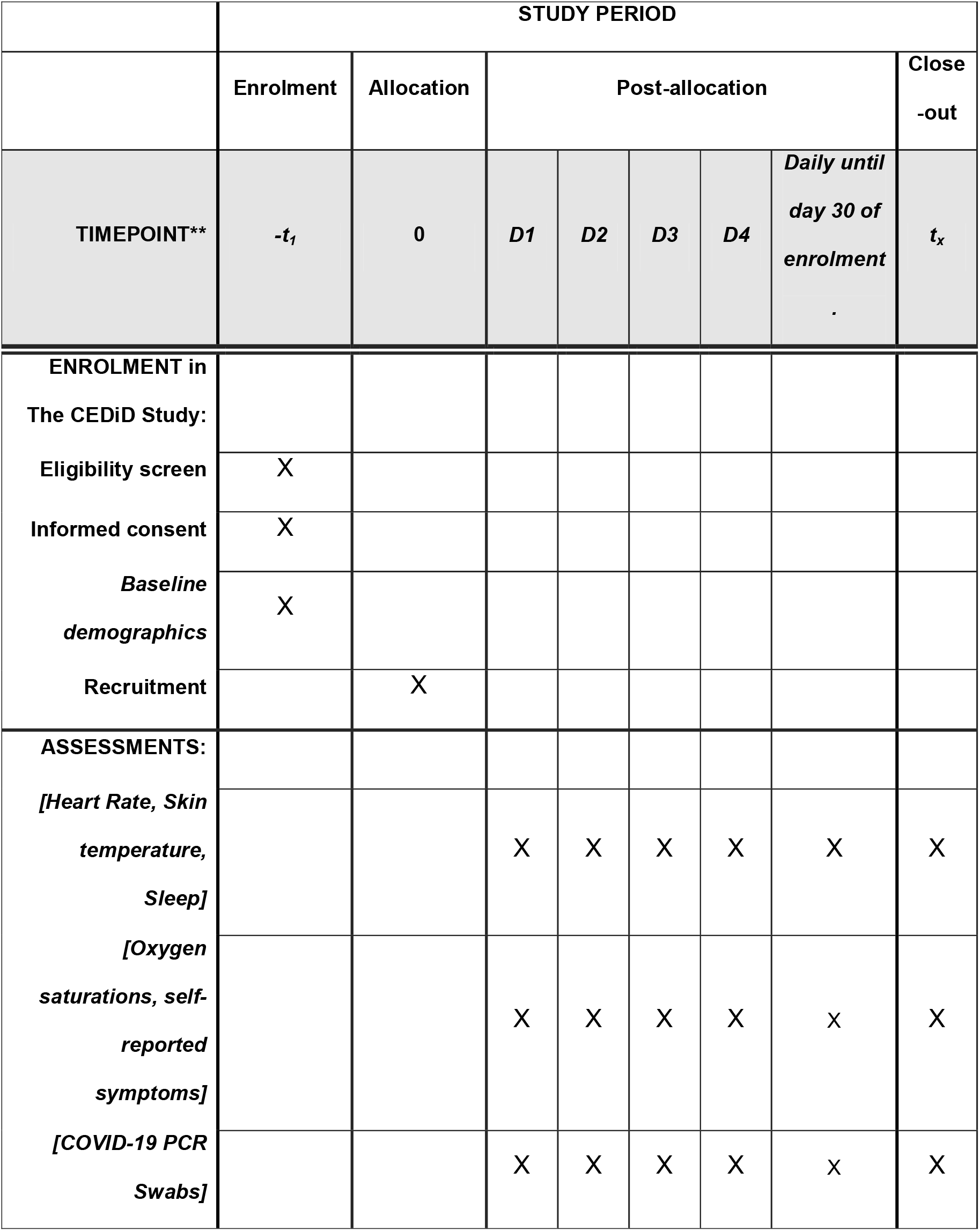
The CEDiD Study (COVID-19 Early Detection in Doctors and Healthcare Workers) SPIRIT Checklist

Following an initial pilot involving a small number of participants in early August, the CEDiD Study will now aim to titrate recruitment in line with predicted future peaks and anticipated caseloads as well as increases in COVID-19-like symptoms in early winter.

### The CEDiD Study

The CEDiD Study is an investigator-initiated single-center prospective observational cohort study in which a total of sixty HCWs at Guy’s and St Thomas’ NHS Foundation Trust will be studied over a thirty day period. A convenience sample of eligible healthcare workers will be taken. Consent will be taken by members of the study team.

The healthcare workers will be given a wearable smart medical device (E4 wristband) from Empatica (Empatica, S.R.L. Via Stendhal 36, 20144 Milan, Italy) which is CE Marked, to wear on their wrists every day during non-working hours for 30 days. The wristband will measure the participant’s temperature, heart rate and sleep status. Participants will also be given a pulse oximeter to measure their oxygen saturations and 30 PCR swabs to self-administer, testing for COVID-19 every day.

Self-reported symptoms will be measured daily via a Microsoft Form attached to a King’s College London server.

The PCR samples will be SARS-CoV-2 swabs (combined nose and throat). They will be self-obtained by HCWs, who will have consented to do this every day for 30 days after reading the informed consent form when enrolling in the study. HCWs will receive a video in which they will be trained how to self-swab appropriately. They will have consented to their samples being processed for COVID-19 detection, viral load, and then frozen for potential use in future research.

Samples will be labelled with participant’s unique study number, appropriately bagged and dropped off in the ward specimen pots. No identifiable information such as name, surname, date of birth, etc. will be included in the label. Where participants are off work for the day they will be asked to keep the samples stored in the appropriate bags in a cool and dry place, before dropping it off when they are next at the hospital.

Samples will then be securely transferred to the King’s College London laboratory (via courier) and according to Government guidelines (10).

Dedicated trained personnel will log reception, store, process and log data from these samples at King’s College London. A unique laboratory number will be assigned to each sample.

Samples will undergo established protocols for SARS-CoV-2 detection (11) which include heat versus no-heat inactivation of samples, nucleic acid extraction using magnetic beads and RT-qPCR employing viral genes and a human gene as a measure of the quality of the sample. Standard curves may be included to determine an estimation of viral copy number present in the samples. Samples will be stored for future analysis including determination of viral titers in cell culture and virus-host responses.

In the event that a participant tests positive for COVID-19 they will be notified via telephone call, advised to self-isolate and contact Occupational Health. Participants will continue to be followed up to monitor their symptoms and self-collected swabs until the end of the trial whenever possible.

### Approvals

The CEDiD Study is approved by the NHS Health Research Authority and Health and Care Research Wales (HCRW) (IRAS Project ID 283321; REC Reference 20/NW/0314) and all necessary local bodies, and was registered at ClinicalTrials.gov (No. NCT04363489)

### Population

The CEDiD Study will include a convenience sample of healthy HCWs who work in high-risk areas for COVID-19. High-risk areas for COVID-19 include wards where HCWs come into direct face-to-face contact with COVID-19 positive patients such as ICU, A&E and COVID positive wards.

Exclusion criteria include HCWs who do not come into face-to-face contact with COVID-19 positive patients, HCWs with previous PCR or antibody positive tests for COVID-19, and HCWs participating in a COVID-19 vaccine trial.

Participants will be followed up on a daily basis via email throughout the study period.

### Exposure

This is an observational cohort study where there will be one group and the cohort of HCWs included will be exposed to COVID-19 through caring for patients who are positive for COVID-19.

### Comparator

A comparator will not be used.

### Outcome Measures in the CEDiD Study

#### Primary outcome measure

The proportion of HCWs who develop COVID-19 infection over the course of the study.

#### Secondary outcome measures

1. The trend in continuous clinical observations during non-working hours
2. The trend in daily-self reported symptoms
3. The development of an algorithm for early detection of COVID-19 in healthcare workers

### Definitions

High-risk areas for COVID-19 are defined as hospital areas where HCWs come into direct face-to-face contact with COVID-19 positive patients on a daily basis. For the purposes of this trial, this setting would include ICU, A&E and COVID-19 positive wards.

HCWs include any healthcare professional who has direct patient contact as part of their daily job in a clinical capacity.

### Data collection and management

Prospectively collected clinical observation data from the E4 wristbands will automatically be uploaded to the secure cloud owned by funders Empatica (with servers located in Virginia, U.S.). Data to be uploaded is physiological data with an ID number for the E4 wristband. The data will be un-identifiable to Empatica as they do not have a link back to an individual. All data will be transferred back to King’s College London/Guy’s and St Thomas’ NHS Foundation Trust, and upon completion of analysis, the data will be deleted from Empatica’s cloud.

Self-reported data including oxygen saturations and daily symptoms will be securely stored on the King’s College London’s system via Microsoft Forms. PCR data will be securely stored on King’s College London’s system. All samples will be labelled with a study ID and no identifying information.

All data will be backed up securely on a hard drive that will not physically leave King’s College London. Only members of the central research team at King’s College London and Guy’s and St Thomas’ NHS Foundation Trust will have access to the participant’s personal data during the study.

Data will be analysed at King’s College London by the central research team. It will be de-identified. Analysis of the swabs’ results will be conducted by the King’s College London laboratory

The following data will be collected in the CEDiD Study at screening: age, sex, previous PCR or antibody positive diagnosis of COVID-19, clinical area of work, comorbidities, current use of medications, and allergies.

The following data will be collected in the CEDiD Study on a daily basis after participants have been enrolled: heart rate, skin temperature, sleep data (via accelerometer), oxygen saturations, symptoms and COVID-19 PCR swabs.

### Safety

Participants will be alerted via telephone if they receive a PCR positive diagnosis of COVID-19, advised to self-isolate and seek advice from occupational health. The protocol includes continued data collection beyond this point to elucidate the pattern of clinical observations. No Significant Adverse Events are expected from the E4 or the oximeters. The E4 wristbands have been tested by Guy’s and St Thomas’ NHS Foundation Trust Medical Physics Department and cleared for safe use.

There is a small chance that participants might experience discomfort/skin abrasion from prolonged use of the wristband without appropriate hygiene practices.

### Statistical analysis

The CEDiD Study data analysis will be performed by the CEDiD Study team after the last participant has finished the study. Time-to-event analysis, descriptive statistics, correlation analysis, hazard ratios and algorithm validation will be performed by the King’s College London statistics team.

### Missing data

In the event of missing data, trends will be used for imputation. Withdrawals will be included in the analysis up to the point of their withdrawal.

### Sample Size

The sample size of the CEDiD Study is set at a minimum of 60 participants in line with established practice for cohort studies collecting both binary and continuous data (12)

### Expected Timeline

July 2020: Approvals obtained and set-up of the CEDiD Study completed

August 2020: Recruitment of a small number of participants to pilot the study

December – January 2021: Expected upscaling of recruitment in view of increased cases of COVID-19 infections within the hospital

February – March 2021: Data analysis, writing and submission of manuscript

### Publication/Authorship

The study manuscript will be completed and submitted to an international, peer-reviewed journal upon completion of the CEDiD Study with results whether positive, negative or neutral. Subsequent analyses of PCR swabs may take place in the future with appropriate consent procedures and approvals. Authors will include named investigators listed in approvals documentation and this manuscript, as well as from King’s College London statistics department.

## Discussion

### Trial Rationale

#### Testing

Testing was reported to be a major factor in the containment of the Ebola outbreak in Africa as it facilitated detection of clusters of infection and appropriate isolation procedures (14). In addition, efficient testing and subsequent contact tracing has been reported as a major enabler of South Korea’s successful containment strategy for COVID-19 (15). Sufficient testing should be carried out, particularly for high-risk groups such as HCWs for SARS-CoV-2, in order to appropriately identify individuals with infection, isolate and contact-trace. On 22^nd^ June 2020, the UK government announced plans for a new saliva test to be administered to the HCWs in Southampton every week as part of a four week pilot (13). Whilst there are emerging methods for testing, nasopharyngeal swabs are the most common types of test used on individuals with mild to moderate disease with a reported sensitivity of 90%, although it should be noted that this is heavily influenced by swabbing technique (16). Participants will be sent a video outlining appropriate swabbing technique.

#### Symptoms

Asymptomatic, or subclinical manifestations of COVID-19 present a considerable challenge to health services which further supports the argument for more regular testing (17,18). There is a variable temporal relationship between exposure positive serology and onset of symptoms, with reports of false-negative swabbing in up to 67% of patients by day five of exposure (19). Within the HCW population, the strongest independent predictors of positive SARS-CoV-2 assays are anosmia, fever and myalgia (20). Whilst there are reliable symptomatic predictors of COVID-19 infection, studies have reported that between 20-50% of the population may be asymptomatic with COVID-19 (21,22). A study of 9,282 HCWs in USA found the most common symptoms amongst HCWs with COVID-19 included a cough (78%), fever (68%), myalgia (64%) and shortness of breath (41%) (4). Amongst HCWs who had an initial negative test, 60% reported a persistent cough (20). There is a paucity of literature exploring other clinical observations such as heart rate and oxygen saturations in HCWs. The CEDiD study will follow-up 60 healthcare workers over 30 days, collecting continuous data on their clinical observations during non-working hours, daily SARS-CoV-2 assays, and daily self-reported symptoms with oxygen saturations to elucidate the trend in these parameters for HCWs working in high-risk areas for COVID-19.

### Strengths and Limitations

In this cohort study, participants will have swabs daily for the duration of the study, in which we will assess the presence of SARS-CoV-2 with the potential of measuring viral titres to provide an insight into the optimum testing strategy. Symptoms and severity can vary depending on factors; including comorbidities, ethnicity, age and sex. As such, the CEDiD Study will seek to include a representative sample.

The limitations include timing as the fluctuations in rate of new infections of COVID-19 are variable on a daily basis. However, recruitment will be of HCWs who are in direct face-to-face contact with COVID positive patients.

If successful, the CEDiD Study will provide important information for healthcare providers in advance of any possible future peaks of infection to prevent spread of disease and ensure minimal disruption to vital healthcare service provision. By leveraging advances in telehealth, medical devices and data analytics, it is hoped that this study will identify new strategies to facilitate the early identification of COVID-19.

### Trial Status

The CEDiD Study started recruitment of participants in August 2020.

## Data Availability

Available upon reasonable request

## List of abbreviations

A&E: Accident and Emergency
CEDiD: COVID-19 Early Detection in Doctors and Healthcare Workers
GSTT: Guy’s and St Thomas’ Hospitals
HCWs: Healthcare Workers
ITU: Intensive Treatment Unit
KCL: King’s College London
PCR: Polymerase Chain Reaction
SARS-CoV-2: Severe Acute Respiratory Syndrome Coronavirus 2

## Declarations

**Consent for publication: N/A**

**Availability of data and materials: Will be made available upon reasonable request to the authors**

## Ethical Approval

The study received HRA and IRAS approval on 16/07/2020 (IRAS Project ID 283321) (REC Reference: 20/NW/0314)

## Guarantor

Dr Kariem El-Boghdadly

## Contributorship

All listed authors have contributed sufficiently in line with ICMJE Authorship Criteria

## Acknowledgements

Thank you to our sponsors King’s College London, co-sponsors Guy’s and St Thomas’ NHS Foundation Trust and funders Empatica for facilitating this clinical trial

